# Identification of B.1.346 lineage of SARS-CoV-2 in Japan: Genomic evidence of re-entry of Clade 20C

**DOI:** 10.1101/2021.01.29.21250798

**Authors:** Kodai Abe, Takako Shimura, Toshiki Takenouchi, Yuka W. Iwasaki, Hirotsugu Ishizu, Yoshifumi Uwamino, Shunsuke Uno, Jun Gotoh, Natsuo Tachikawa, Yuriko Takeuchi, Junpei Katayama, Hiroyuki Nozaki, Susumu Fujii, Shikou Seki, Morio Nakamura, Kazuhiro Uda, Takahiko Misumi, Jun Ishihara, Kenichiro Yamada, Toshio Kanai, Shinji Murai, Kazuhiro Araki, Tamotsu Ebihara, Haruhiko Shiomi, Naoki Hasegawa, Yuko Kitagawa, Masayuki Amagai, Makoto Suematsu, Kenjiro Kosaki

## Abstract

**Objectives:** Whole SARS-CoV-2 genome sequencing from COVID-19 patients is useful for infection control and regional trends evaluation. We report a lineage data collected from hospitals in the Kanto region of Japan.

**Methods:** We performed whole genome sequencing in specimens of 198 COVID-19 patients at 13 collaborating hospitals in the Kanto region. Phylogenetic analysis and fingerprinting of the nucleotide substitutions underwent to differentiate and classify the viral lineages.

**Results:** More than 90% of the strains belonged to Clade 20B and two lineages (B.1.1.284 and B.1.1.214) have been detected predominantly in the Kanto region. However, one sample from a COVID-19 patient in November 2020, belonged to the B.1.346 lineage of Clade 20C, which has been prevalent in western United States. The patient had no history of overseas travel and no contact with anyone who had travelled abroad, suggesting that this strain appeared likely to have been imported from western United States, across the strict quarantine barrier.

**Conclusion:** B.1.1.284 and B.1.1.214 have been identified predominantly in the Kanto region and B.1.346 of clade 20C in one patient was probably imported from western United States. These results illustrate that a decentralized network of hospitals can be significantly advantageous for monitoring regional molecular epidemiologic trends.

**Highlights:**

· Whole SARS-CoV-2 genome sequencing is useful for infection control
· B.1.1.284 and B.1.1.214 have been identified predominantly in the Kanto region
· B.1.346 of Clade 20C was detected in one COVID-19 patient in November
· Molecular genomic data sharing provides benefits to public health against COVID-19

## Introduction

Whole SARS-CoV-2 genome sequencing of samples from COVID19 patients is useful for infection control (Takenouchi T, et al., 2020). When multiple COVID19-positive cases occur simultaneously in a hospital, it is critical to determine whether newly diagnosed patients have nosocomial infection or community infection. Under nosocomial infection, a thorough contact tracing of healthcare workers and inpatients is essential whereas in the situation of community infection, such a persisted intrahospital surveillance may not be necessary.

Datasets of whole viral genome assembled from multiple hospitals can give critical clues on regional or national trends. National surveillance system developed in the United Kingdom (UK) clarified that more than 1000 lineages spread during pre-lockdown high travel volumes and few restrictions on international travel (Du Plessis L, et al., 2021). Recently, the genomic surveillance system successfully identified two variants of concern, 501Y.V1 (B.1.1.7 originated from the UK) and 501Y.V2 (B.1.351 originated from South Africa), both of which spread rather quickly (Davies N, et al., 2020; European Centre for Disease Prevention and Control, 2020). In Japan, eight laboratories including our group have deposited whole viral genome data to the public sequence database GISAID (https://www.gisaid.org/): A total of 9943 sequences have been deposited and some results of national survey have been summarized (Sekizuka, et al., 2020; Sekizuka, et al., 2020). Despite the requirement by international convention, regional origin of the samples or precise date of sampling has not been specified in most of the publicly available data derived from Japan.

Here, we report the lineage summary of our data collected from 13 hospitals located in the Kanto region of Japan to document relatively recent trends in and around the Tokyo Metropolitan area. Whole genome data from 198 specimens which were accumulated and utilized for infection control showed that the lineages of the strains were limited to only two closely related strains, B.1.1.284 and B.1.1.214. The observation supports success of quarantine system. However, one strain derived from a patient with nosocomial COVID-19 infection belonged to the B.1.346 lineage which is prevalent in the western United States (USA) as of winter 2020. We suspect that the episode is attributed to undetected imported infections at the border.

## Materials and Methods

### Study design and patients

The study protocol was approved by the Institutional Review Board of Keio University School of Medicine (approval number: 20200062) with each collaborating hospitals’ review board and was conducted according to the principles of the Declaration of Helsinki.

### DNA sequencing method

Whole viral genome sequence was determined as previously reported (Takenouchi et al., 2020). PCR-based amplification was performed using the Artic ncov-2019 primers in addition to a No.72-mutant primer, version 3 (https://github.com/artic-network/artic-ncov2019/blob/master/primer_schemes/nCoV-2019/V3/nCoV-2019.tsv) in two multiplex reactions according to the globally accepted “nCoV-2019 sequencing protocol” (https://www.protocols.io/view/ncov-2019-sequencingprotocol-bbmuik6w). The sequencing library for amplicon sequencing was prepared using the Next Ultra II DNA Library Prep Kit for Illumina (New England Biolabs). Paired end sequencing was performed on the MiSeq platform (Illumina, CA). The bioinformatic pipeline used in this study, “Mutation calling pipeline for amplicon-based sequencing of the SARS-CoV-2 viral genome,” is available at https://cmg.med.keio.ac.jp/sars-cov-2/. All single nucleotide substitutions including non-synonymous and synonymous mutations were annotated by the ANNOVAR software and were assessed using VarSifter (https://research.nhgri.nih.gov/software/VarSifter/). The analytic protocol is described previously in our publication, submitted in November 24, 2020, in preprint server medRxiv (Abe, et al., 2020).

### Genetic clade or lineage naming in phylogenic tree analyses

A phylogenetic tree analysis was performed locally using the Augur program available from Nextstrain (https://nextstrain.org/) and genome sequence data obtained in the currently reported study as well as data available from the global database EpiCov hosted at GISAID (Hadfield, et al., 2020). The Nextclade (https://clades.nextstrain.org/) was also used to generate fingerprinting patterns to visualize the SARS-CoV-2 sequence alignments and similarities/identities among samples.

We used the international genetic clade nomenclature system defined by Nextstrain.org (https://virological.org/t/updated-nextstain-sars-cov-2-clade-naming-strategy/581) (Figure 1). We also used the software Phylogenetic Assignment of Named Global Outbreak Lineages (Pangolin; https://cov-lineages.org/index.html) to assign viral lineages in an automatic and precise manner (Rambaut, et al., 2020).

**Figure 1.**
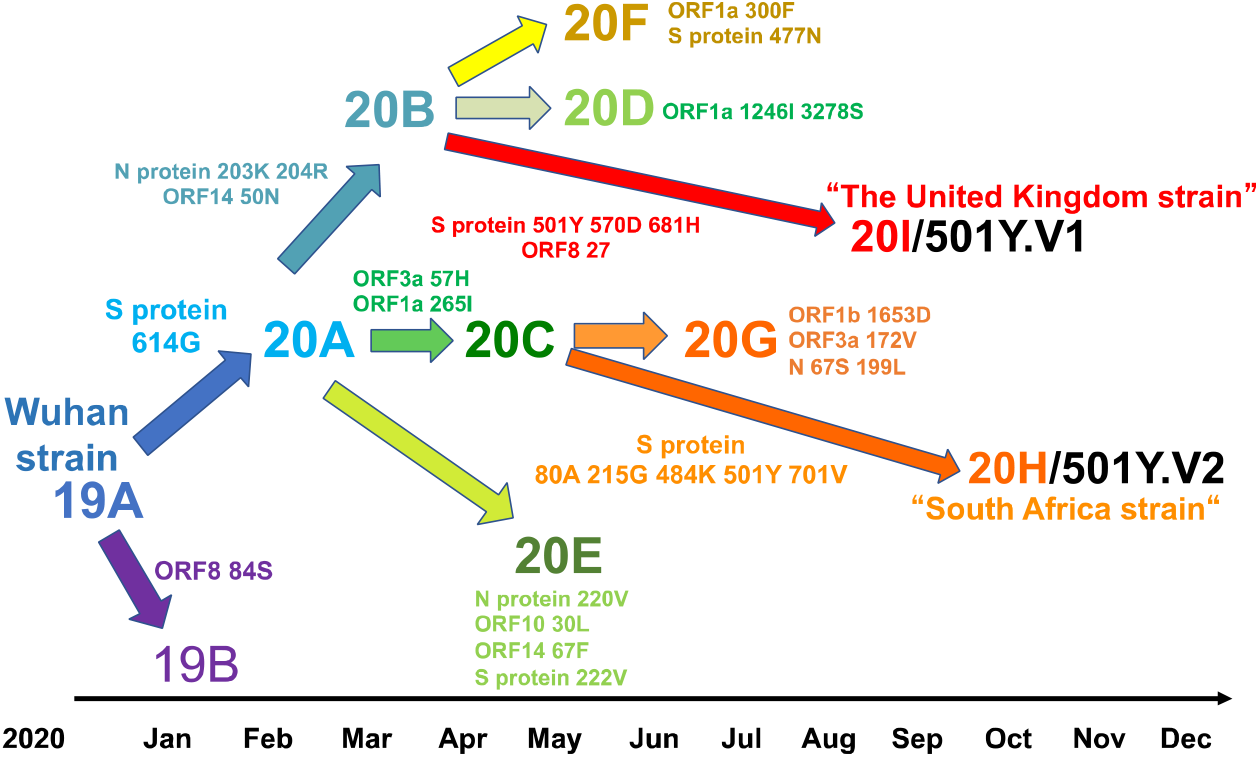
International genetic clade defined by Nextstrain.org. The Wuhan strain was defined as 19A, with a large clade derived dendritically. The currently prevalent British mutant strain was defined as 20I (501Y.V1) and South Africa as 20H (501Y.V2).

## Results

Of the 198 strains, 189 (95.5%) belonged to Clade 20B of the Clade system. Ninety-eight of 189 (51.9%) were classified as B.1.1.284 (referred to as ‘Clade 20B-T’ in our previous publication; Abe et al., 2020) whereas 60 (31.7%) were classified as B.1.1.214. These two lineages differ by six nucleotide substitutions. According to the GISAID database, the total number of samples corresponding to B.1.1.284 and B.1.214 are 4183 and 1070. Vast majority of B.1.1.284 and B.1.214 derived from Japan. Outside Japan, B.1.1.284 and B.1.1.214 are extremely rare. Only 8 B.1.1.284 samples have been detected (2 in Thailand, 2 in South Korea, 2 in Australia, 1 in Singapore, and 1 in Hongkong. Only 3 B.1.1.214 samples have been detected (2 in Australia and 1 in the USA). The other lineages of Clade 20B were also detected; 15 in B.1.1.285 lineage, 10 in B.1.1.114 lineage, 4 in B.1.1.119, and 1 in B.1.1.163.

One strain of the B.1.346 lineage belonging to clade 20C that likely emerged from New York (Zhang, et al., 2021) was detected in one patient with confirmed COVID-19 infection at a hospital in the Kanto region in November 2020 (Figure 2). The patient did not have any history of overseas travel or known contact with anyone who had travelled abroad. The strain had 18 single nucleotide substitutions in comparison with the original Wuhan SARS-CoV-2 sequence (ID. NC_045512.2). Functional relevance of the single nucleotide substitutions is summarized in Table 1.

**Table 1.** Single nucleotide substitution and functional relevance of B.1.346 in 20C compared with that of Wuhan strain

| Position in reference to the Wuhan strain | Reference allele | Variant allele | Protein | Amino acid change | annotation | Note |
| --- | --- | --- | --- | --- | --- | --- |
| 241 | C | T | 5'UTR | NA | NA |  |
| 1059 | C | T | NSP2 | T85I | Non-Structural protein 2 | 20C signature |
| 3037 | C | T | NSP3 | F106F | Predicted phosphoesterase, papain-like proteinase | 20A signature |
| 6948 | A | C | NSP3 | N1410T | Predicted phosphoesterase, papain-like proteinase | Common among 5 strains from the western USA |
| 12820 | A | G | NSP9 | L45L | ssRNA-binding protein |  |
| 14408 | C | T | NSP12b | P314L | RNA-dependent RNA polymerase, post-ribosomal frameshift | 20A signature |
| 14587 | G | T | NSP12b | A374S | RNA-dependent RNA polymerase, post-ribosomal frameshift |  |
| 15324 | C | T | NSP12b | N619N | RNA-dependent RNA polymerase, post-ribosomal frameshift | Common among 5 strains from the western USA |
| 15403 | T | C | NSP12b | R645R | RNA-dependent RNA polymerase, post-ribosomal frameshift | Common among 5 strains from the western USA |
| 16762 | C | T | NSP13 | L176F | Helicase |  |
| 17637 | A | G | NSP13 | K467K | Helicase |  |
| 17880 | A | G | NSP13 | Q548Q | Helicase |  |
| 20762 | C | T | NSP16 | T35I | 2'-O-ribose methyltransferase |  |
| 23403 | A | G | S | D614G | Spike | 20A signature |
| 24337 | C | T | S | N925N | Spike | Common among 5 strains from the western USA |
| 25563 | G | T | ORF3a | Q57H | ORF3a protein | 20C signature |
| 26735 | C | T | M | Y71Y | Membrane | Common among 4 strains from the western USA |
| 28887 | C | T | N | T205I | Nucleocapsid protein |  |

NSP, non-structural polyprotein; ORF, open reading frame; ssRNA, single-strand RNA; NA, not assessed.

**Figure 2.**
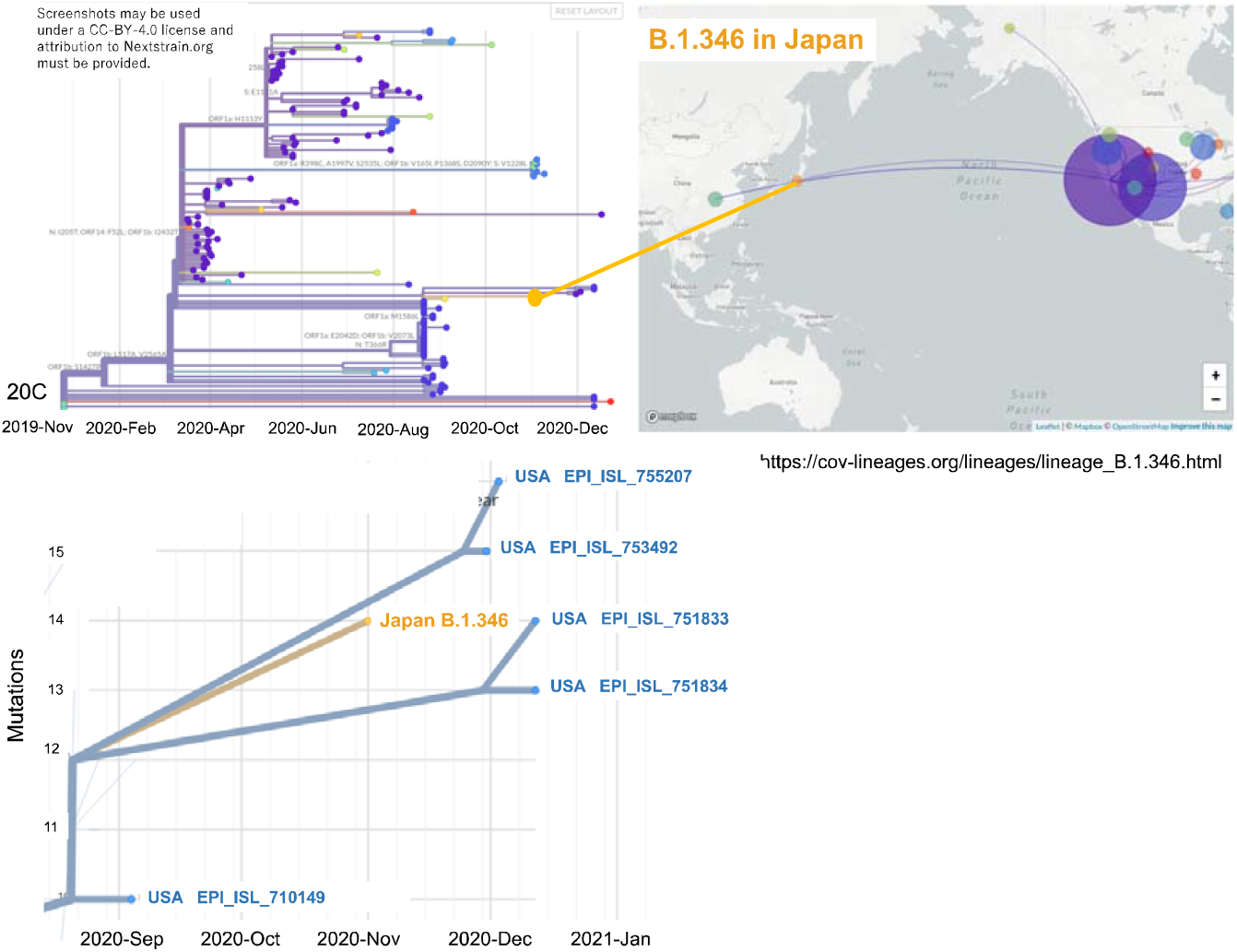
Relationship between Japan and the World in the Clade 20C. Internationally, more than 90% of Clade 20C has been detected from the USA. The whole genome sequence of a sample collected from one COVID-19 positive patient at a hospital in the Kanto area at the end of November 2020 was found to be very similar to a virus strain prevalent on the West States of the United States (a type of 20C, B.1.346) according to phylogenetic tree analysis.

According to the GISAID database, 159 strains belonging to the B.1.346 lineage have been deposited in the database (acknowledgment table). Most strains derived from the western United States. Detailed examination of the phylogenetic tree revealed that 4 strains were very closely related (Figure 2). The genomic data strongly suggest that this strain was presumably imported into Japan across the quarantine barrier. The GISAID data from Japan indicate that the last clade 20C strains detected in Japan were in March and May, 2020, but not thereafter, except in quarantine cases, supporting the notion that the newly identified clade 20C strain in this study was indeed imported from overseas.

Comparison of the fingerprinting pattern of the B.1.346 lineage strains detected in Japan in this study and those detected in the western United States and a representative clade 20C strain detected in Japan in the spring of 2020 (Figure 3) further augments the notion that the strain of B.1.346 lineage belonging to clade 20C was imported into Japan across the quarantine barrier from western United States.

**Figure 3.**
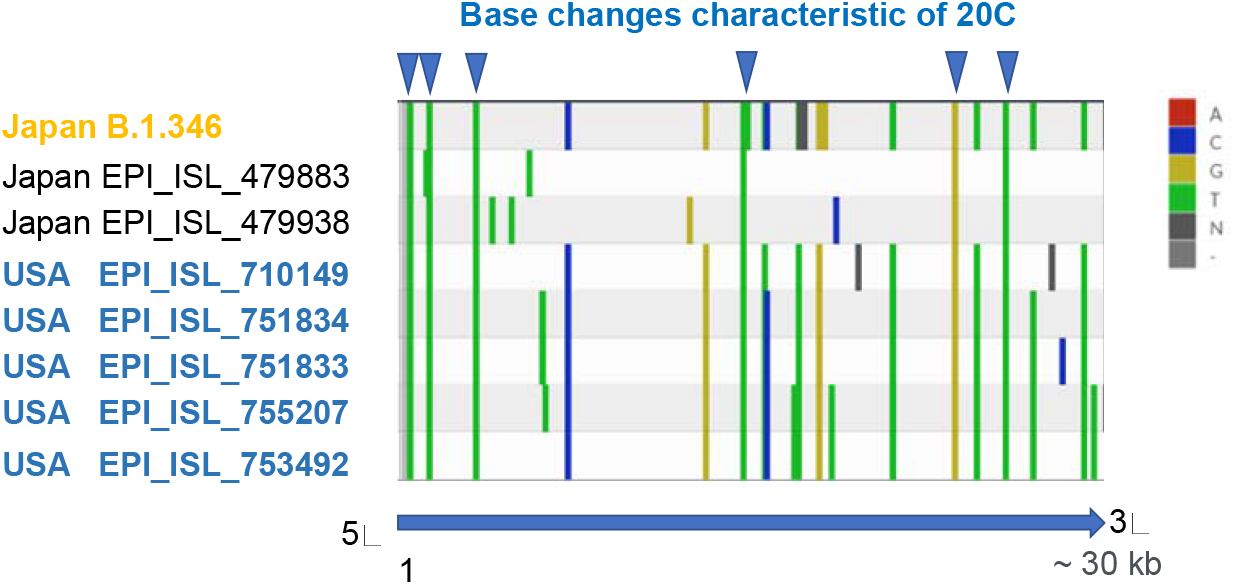
The Nextclade analysis of B.1.346 Finger printing diagram with past Japanese 20C strains. The B.1.346 strain found in this study in November 2020 is very similar to the five strains from the West States of the United States in terms of mutation sites, while compared to the strains belonging to Clade 20C found in domestic quarantine around May, the majority of the strains have different mutation sites, although there are some similarities.

There is no evidence that the newly detected clade 20C strain in the Kanto region shows altered transmissibility or virulence, unlike variants of concern such as 501Y.V1 (B.1.1.7) in the UK and 501Y.V2 (B.1.351) in South Africa.

## Discussion

Through lineage analysis of 198 whole viral genome data accumulated from 13 hospitals in the Kanto region, we have herein concluded the following: First, most of the strains belong to B.1.1.284 and B.1.1.214. Second, at least one strain was imported from the USA.

The fact that only two strains have been detected predominantly in the Kanto region supports a notion that Japan’s quarantine system had relatively been successful after national lock down in April and May of 2020. The observation that extremely limited number of lineages (i.e., 2) makes sharp contrast to that in the UK and other countries where numerous numbers of strains are still prevailing (Popa, et al., 2020; Lemiux, et al., 2020). In view of emergence of potentially virulent strains (501Y) in the UK and South Africa, thorough and strict quarantine system should further be warranted in Japan.

The implication of detecting viral strain prevalent in West States of the United States is two folds. First, no quarantine policy can be perfect. If the policy is reset to be less strict, close follow-up genomic monitoring is mandatory. Because of the internationally unprecedent uniformness of the domestic viral strains in Japan, detection of foreign strains may be relatively easy as exemplified in this single patient. Second, if entry of foreign strain does occur in the near future, it may be relatively easy to pinpoint the geographic origin of the incoming strain by the number of nucleotide substitutions in the SARS-CoV-2 genome.

Overall, the viral genomic monitoring system for in-hospital infections reported herein enabled us to unravel regional or national trends with agility. Until now in Japan, a national centralized network system, composed of National Institute of Infectious Diseases in Tokyo and Public Health Centers and Public Health Institutes had been put in place for the purpose of viral genome surveillance. Our report illustrates that a decentralized network system composed of multiple hospitals mutually sharing molecular genomic data in near real-time provides robust benefits to public health with agility, under the conditions of this COVID-19 pandemic. Molecular genomic data obtained through such a system could be swiftly reflected in the national decision-making process for public health practices, including the strictness of quarantine measures or implementation of lockdowns in response to the detection of SARS-CoV-2 variants of concern.

## Supporting information

Acknowledgment table

## Data Availability

Data sharing
We downloaded the full nucleotide sequences of the SARS-CoV-2 genomes from the GISAID database (https://www.gisaid.org/). A table of the contributors is available (acknowledgment table). We have uploaded the full nucleotide sequences of our cohort to the GISAID database.

## Author contributions

KAb and TS contributed to writing of the report and data analysis. YI, HI, TT and HS contributed to sequencing and analysis. YU, SU, JG, NT, YT, JK, HN, SF, SS, MN and KU contributed to clinical data and clinical care. TM, JI, KY, TK, SM, KAr and TE contributed to public health intelligence and case identification. NH, YK and MA contributed to writing and editing of the report. KK had the original idea for the study and contributed to diagnostics, formal analysis, and writing and editing of the report.

## Acknowledgements

We thank all the patients and healthcare workers who have fought against COVID-19. This work was supported by the Keio Donner Project and is devoted to the late Professor Shibasaburo Kitasato, the founder of Keio University School of Medicine. We also thank SRL, Inc.

## Data sharing

We downloaded the full nucleotide sequences of the SARS-CoV-2 genomes from the GISAID database (https://www.gisaid.org/). A table of the contributors is available (acknowledgment table). We have uploaded the full nucleotide sequences of our cohort to the GISAID database.

